# Study the combination of brain MRI imaging and other datatypes to improve Alzheimer’s disease diagnosis

**DOI:** 10.1101/2022.10.30.22281735

**Authors:** Jonathan Stubblefield, Alan Kronberger, Jason Causey, Jake Qualls, Jennifer Fowler, Kaiman Zeng, Karl Walker, Xiuzhen Huang, the Alzheimer’s Disease Neuroimaging Initiative

## Abstract

Alzheimer’s Disease (AD) is a degenerative brain disease and is the most common cause of dementia. Despite being a common disease, AD is poorly understood. Current medical treatments for AD are aimed at slowing the progression of the disease. So early detection of AD is important to intervene at an early stage of the disease. In recent years, by using machine learning predictive algorithms, assisted clinic diagnosis has received great attention due to its success of machine learning advances in the domains of computer vision. In this study, we have combined brain MRI imaging features and the features of other datatypes, and adopted various models, including XGBoost, logistic regression, and k-Nearest Neighbors, to improve AD diagnosis. We evaluated the models on the benchmark dataset of Alzheimer’s Disease Neuroimaging Initiative. Experiment results show that the logistic regression model is the top performer in terms of evaluation metrics of precision, recall, and F1-score. The prediction of the models could provide valuable information for diagnosis and prognosis of patients with suspected Alzheimer’s disease. The XGBoost model achieves a comparable performance and has the potential to serve as a valuable diagnostic tool for patients with suspected AD with its self-validation by rediscovering previously known associations with AD.

## 1. Background

Alzheimer’s disease (AD) is a degenerative brain disease and is the number one cause of dementia [@Wolk]. It is estimated that 60% - 80% of dementia cases are due to AD or have shown Alzheimer’s disease brain changes with one or more other causes of dementia, such as Cerebrovascular disease or Lewy body disease [@ADFact&Figure]. AD typically manifests with impairment in memory, judgement, executive function, cognitive decline, and behavioral and psychological changes [@Wolk]. Despite being a common disease, AD is poorly understood [@Wolk]. The cause and pathogenesis of AD remain open areas of research [@Wolk]. As of yet, there is no cure for AD [@Wolk]. Current medical treatments for AD are aimed at slowing the progression of the disease [@Wolk].

### 1.1 Current Diagnostics

The only definitive diagnosis for AD is a brain biopsy with a histopathological examination, which is usually only done during an autopsy [@Wolk]. This makes the diagnosis of AD very challenging. In practice, AD is usually diagnoses by a physician’s clinical assessment of the patient [@Wolk]. Neuroimaging, such as Magnetic Resonance Imaging (MRI) and Positron Emission Tomography (PET), can be used to search for alternative diagnoses or document the progression of the disease [@Wolk]. Biomarkers for AD have been studied and can support a diagnosis of AD but are not currently recommended as diagnostic tests [@Wolk].

In recent years, data-driven approaches have received great attention due to the advancement of big data, computational power, and machine learning algorithms. Because the AD resulted brain changes is a slowly developing progress and could begin years before symptoms emerge [@ADFact&Figure], early detection of AD has a great potential to provide early intervention, slow disease progression, and reduce the care cost.

### 1.2 Related Work

Machine learning algorithms have been applied in the medical field to assist diagnosis for a lot of diseases, such as heart disease, Parkinson’s disease, and Coronavirus disease. A great amount of research has been performed on using machine learning algorithms to assist AD diagnosis, which trains a classification model to predict if a patient will have AD, develop mild cognitive impairment (MRI), or remain normal (represented as normal control, NC). The studied machine learning algorithms can be categorized as classical machine learning models and deep learning models. The classical machine learning models mainly include logistic regression, support vector machine (SVM), and tree-based models. The deep learning models are different architectures of neural networks.

Antor et. al. performed a comparative analysis of logistic regression, decision tree, random forest, and SVM models to predict AD. They also evaluated the models with fine-tuning to those without fine-tune. Their experiment results showed that the SVM best performed in detecting dementia in terms of accuracy, recall, precision, AUC, and F1 score. They claimed a more complex model such as random forest classifier suffered from overfitting.

Grueso and Viejo-Sobera [@ML systematic review] conducted a comprehensive review and qualitative analysis over 116 studies selected from 452 publications that used machine learning to predict progression of AD. It was found that the SVM was the most commonly used a classifier with a mean accuracy of 75.4%. The SVM algorithm uses a kernel function to map the feature space to a higher dimension and usually a polynomial or Gaussion function was used [@Automated]. Ensembling models of SVM and other methods have also been proposed to optimize the feature selection and to improve the generalization of models [@Thung].

Deep learning techniques have also gained great attentions in recent years due to its success in the domains of computer vision and natural language processing. Saleem [@DeepLearning] et. al. have reviewed a number of studies for AD diagnosis based on the major deep learning models, including feed-forward deep neural networks (DNN), convolutional neural networks (CNN), auto-encoder (AE), recurrent neural networks (RNN), deep belief network (DBN), generative adversarial networks (RNN), and hybrid models. The authors observed deep learning techniques in general outperform classical machine learning techniques, with accuracies ranging from 53.17% to 99.95% for classifications of NC/AD, NC/AD, MCI/AD or NC/MCI/AD. However, most deep learning models lack explainability and as a result lead to criticisms for being adopted into clinical practice. Paper [@Explainability] discussed the role of explainable AI from the perspectives of technology, medicine, legislation, ethics, and societies.

They argued that emitting explainability in clinical decision system would jeopardize the core ethic value of medicine and public health.

### 1.3 Alzheimer’s Disease Neuroimaging Initiative (ADNI)

To combat this current lack of usable diagnostic studies the ADNI study was started in 2004 [@about_adni]. This large, longitudinal, multi-center study has accumulated data on a large number of patients with data clinical, MRI imaging, and proteomic data [@about_adni].

## 2. Methods

Data used in the preparation of this article were obtained from the Alzheimer’s Disease Neuroimaging Initiative (ADNI) database (adni.loni.usc.edu). The ADNI was launched in 2003 as a public-private partnership, led by Principal Investigator Michael W. Weiner, MD. The primary goal of ADNI has been to test whether serial magnetic resonance imaging (MRI), positron emission tomography (PET), other biological markers, and clinical and neuropsychological assessment can be combined to measure the progression of mild cognitive impairment (MCI) and early Alzheimer’s disease (AD).

In this paper, we present a set of models developed with data from the ADNI study for the diagnosis of AD using proteomic and MRI imaging data.

### 2.1 ADNI Patients

We used the ADNI dataset for this study. We sampled 167 individuals across the ADNI 1 and ADNI 2 datasets that had both proteomic and MRI imaging data. These individuals included a mix of patients with AD, mild-cognitive impairment, and healthy controls.

### 2.2 ADNI Imaging Features

We used the ADNI imaging features that had already been processed by the study using FreeSurfer. FreeSurfer is an open-source software package used to visualize and analyze MRI images. The MRI data of the ADNI 1 dataset was obtained using FS V4.4. FS V5.1 was used to process the ADNI 2 dataset [@adni_methods]. The results contained 70 cortical thickness measurements and 42 cortical volume measurements [@adni_methods].

### 2.3 ADNI Proteomic Features

The ADNI proteomic features consist of protein levels from samples of the patients’ cerebrospinal fluid. These samples were processed by Rules-Based Medicine using the Luminex xMAP [@biomarkers]. The data contained a total of 229 proteomic measurements.

### 2.4 Feature Normalization

All features were scaled element-wise using Scikit-Learn version 0.24.1’s MinMaxScaler [@Pedregosa].

### 2.5 Different methods

#### XGBoost Method

We used XGBoost as our classification model during this project [@Chen]. We recorded results for each pairwise binary classification problem for the three classes, normal control (NC), mild-cognitive impairment (MCI), and Alzheimer’s Disease (AD). We also recorded results for XGBoost classifying all three classes simultaneously. We ran the model in Python using Python 3.8.8 and the 1.3.3 version of the XGBoost package. The experiment was performed using 5-fold cross-validation with Scikit-Learn’s stratified k-fold [@Pedregosa]. We used the 0.24.1 version of Scikit-Learn [@Pedregosa]. XGBoost’s performance was evaluated on each datatype individually and both data types combined. The two datatypes were combined using simple concatenation of the feature arrays.

#### Logistic Regression Method

Logistic regression was chosen as a comparison model to XGBoost. Functioning as linear model predicting class probabilities, we expected it to approach the problem in very different than XGBoost’s gradient boosted trees [@Chen]. We used Scikit-Learn version 0.24.1’s version of logistic regression with default parameters aside from a max iteration of 500 [@Pedregosa].

#### K-Nearest Neighbors Method

K-Nearest neighbors (KNN) was also chosen as a comparison model. KNN is a non-parametric model that can vary its flexibility by changing the value of K. We used Sciki-Learn version 0.24.1’s version of KNN, evaluating it for K values 2, 5, and 15 [@Pedregosa].

#### Model Evaluation

Each method was evaluated using five-fold cross-validation on each binary classification problem consisting of two paired classes (control vs Alzheimer’s, control vs mild cognitive impairment, and mind cognitive impairment vs Alzheimer’s) as well as the 3-class classification problem. Recall, precision, and f1-score were calculated for each classification problem and each fold. The averages with respect to each class were reported.

#### SHAP Feature Importance

We used the SHAP TreeExplainer utility to evaluate and graph the relative importance of the imaging and proteomic features to the XGBoost model [@Lundberg_tree].

## 3. Results

Here we present the performance of the different methods with the MRI image features and proteomics data features respectively, and the combination of the two types of data features.

**Figure 1:**
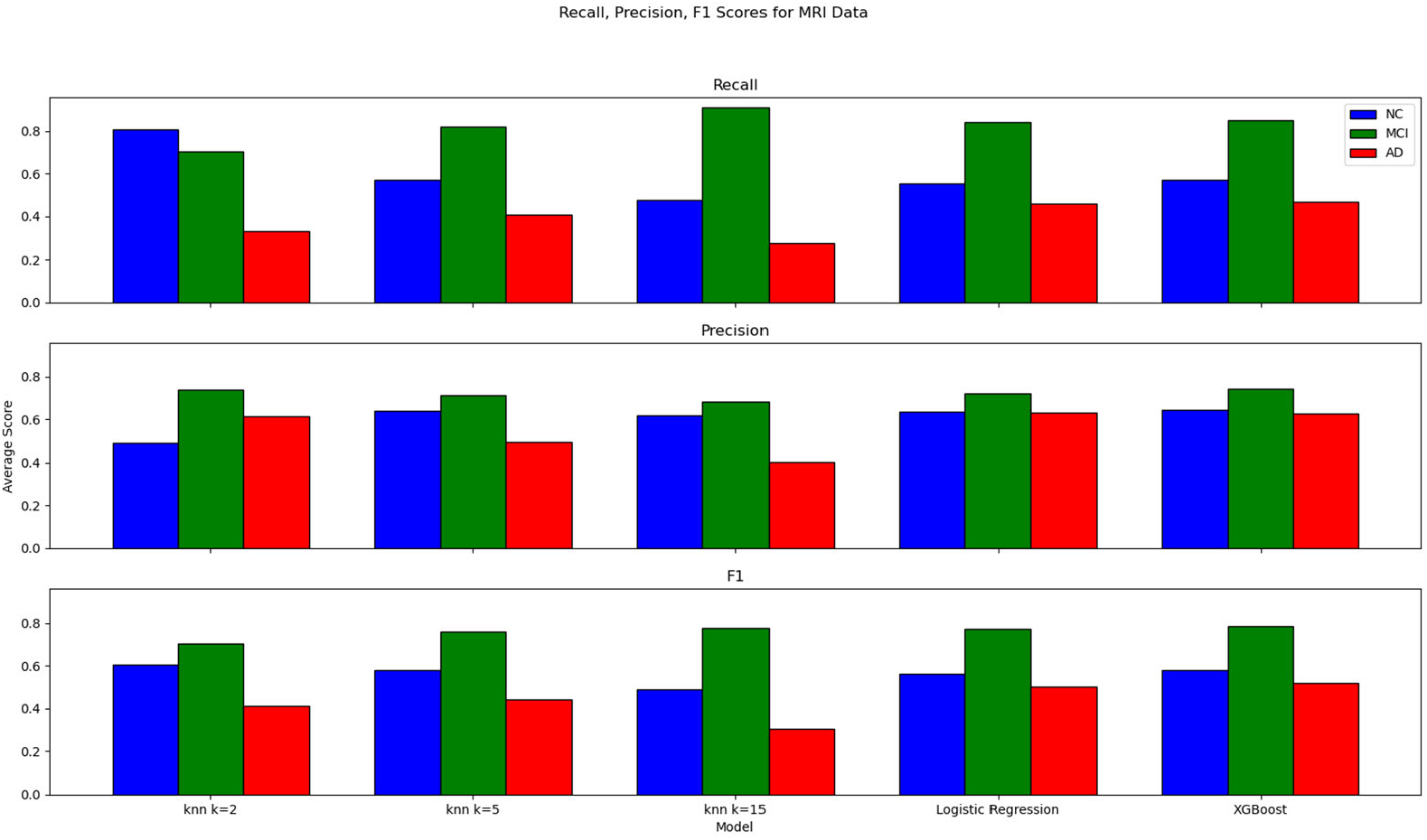
Recall, Precision, F1 Scores for MRI Data. These graphs show various classification metrics with respect to each class. From top to bottom, the graphs show recall, precision, and f1-score. Metrics for the NC class are displayed in blue, MCI in green, and AD in red. The metrics are the averages across all trials.

**Figure 2:**
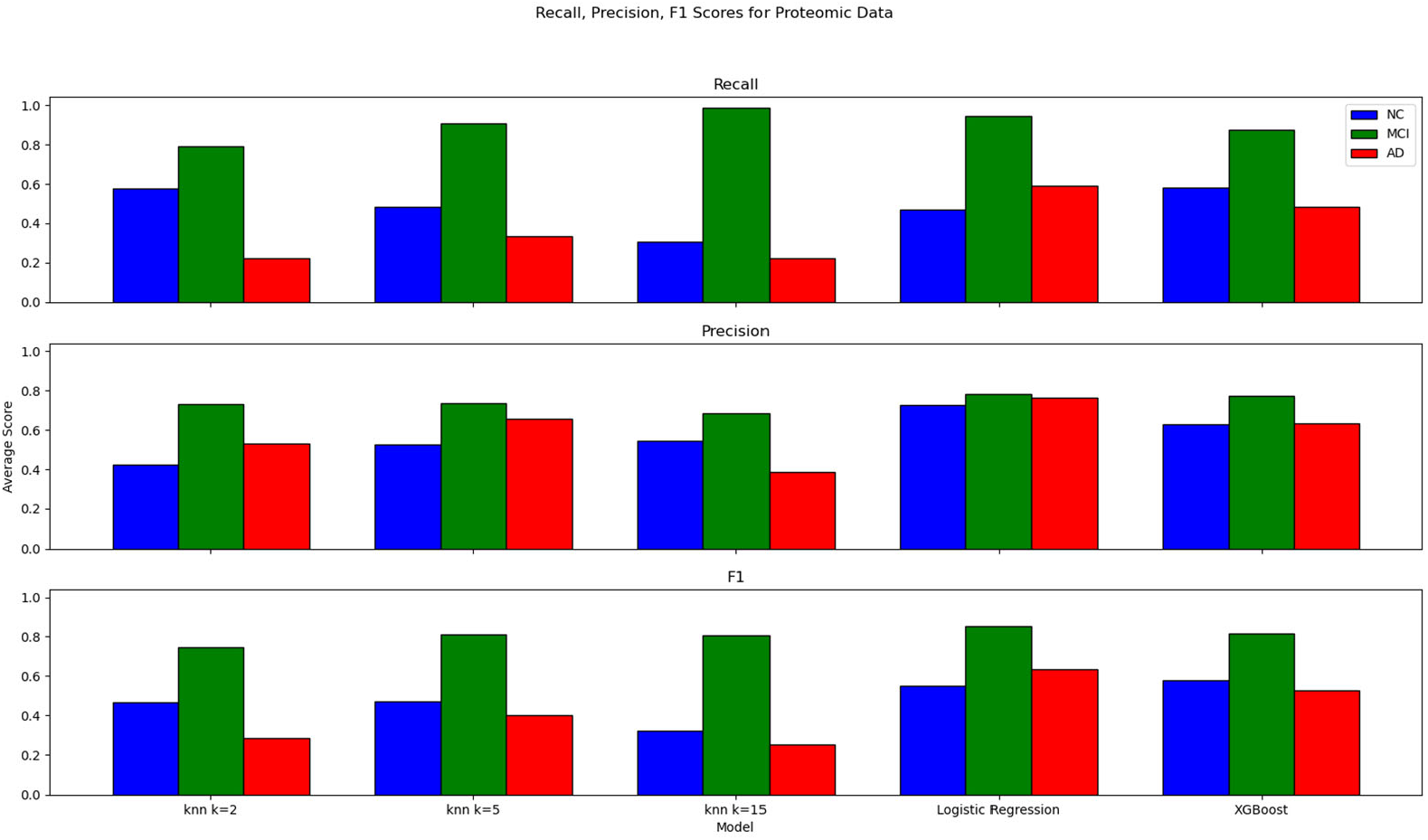
Recall, Precision, F1 Scores for Protein Data. These graphs show various classification metrics with respect to each class. From top to bottom, the graphs show recall, precision, and f1-score. Metrics for the NC class are displayed in blue, MCI in green, and AD in red. These metrics are the averages across all trials.

**Figure 3:**
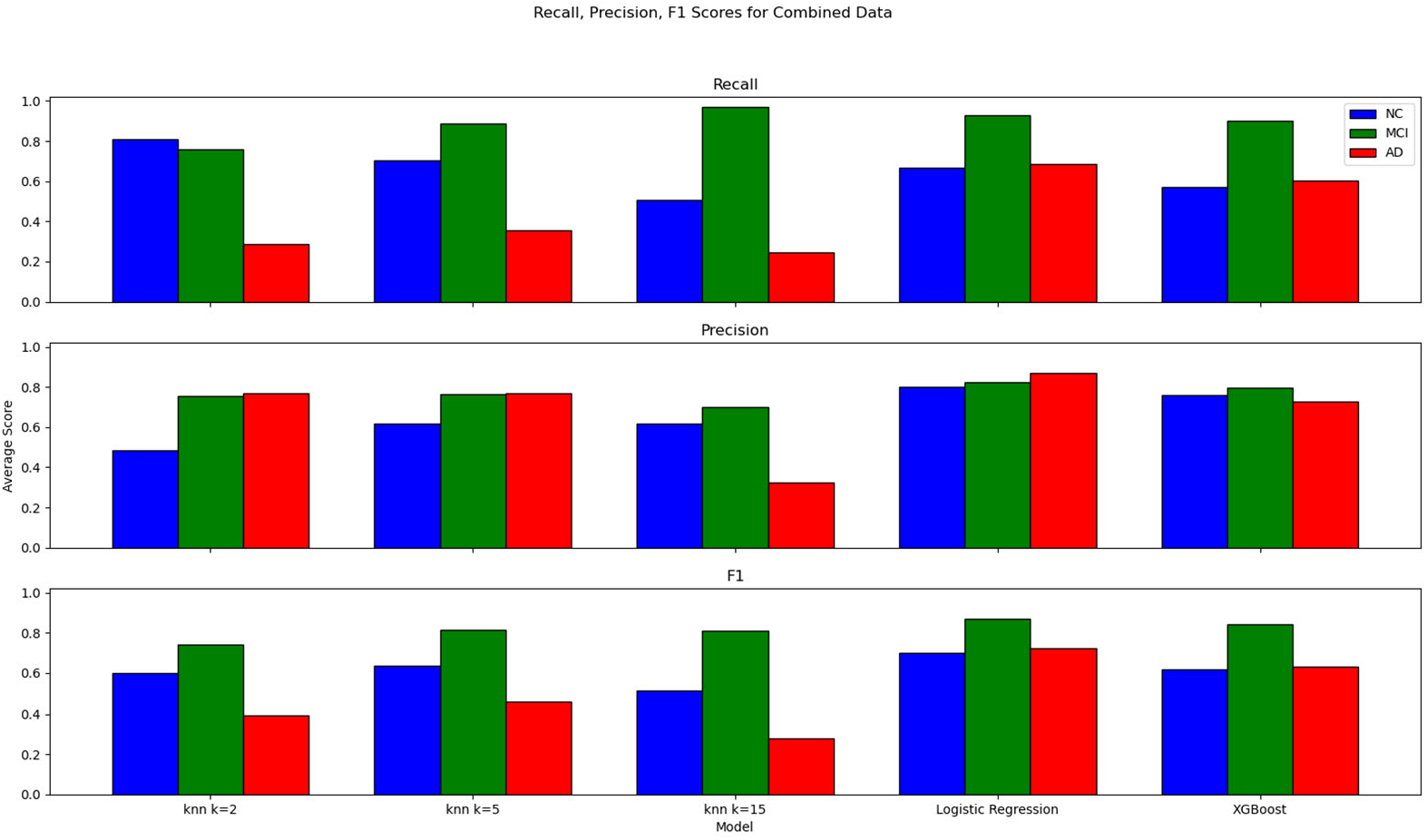
Recall, Precision, F1 Scores for Combined Data. These graphs show various classification metrics with respect to each class. From top to bottom, the graphs show recall, precision, and f1-score. Metrics for the NC class are displayed in blue, MCI in green, and AD in red. These metrics are the averages across all trials.

**Figure 4:**
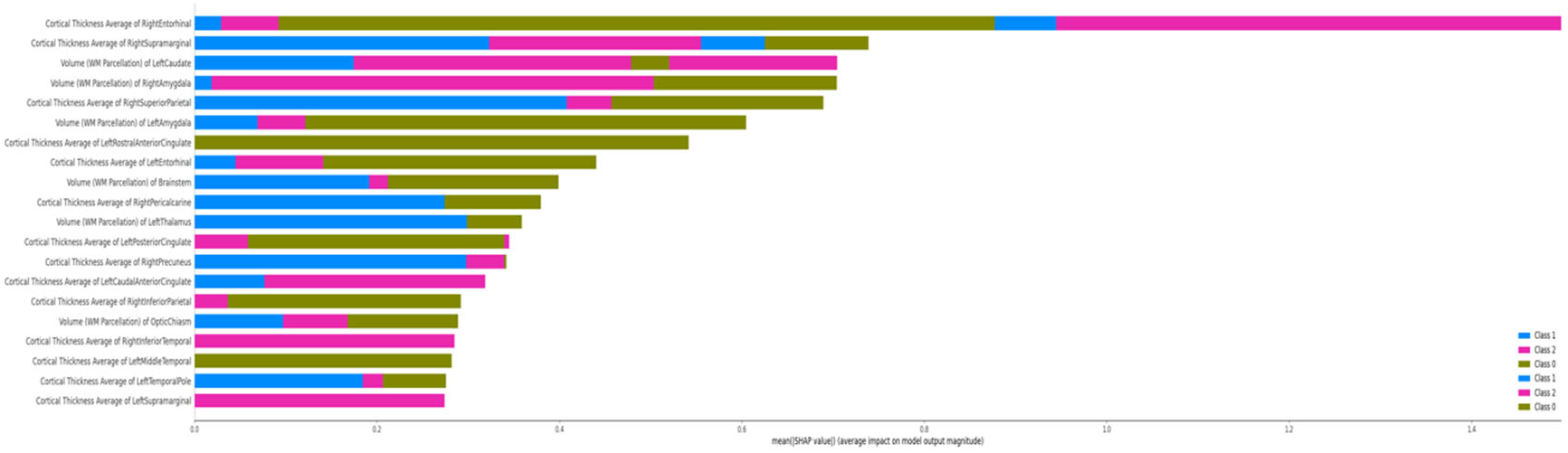
ADNI Summary Plot, Image Features. The overlapping bars show the average SHAP values of the top image features for each class.

**Figure 5:**
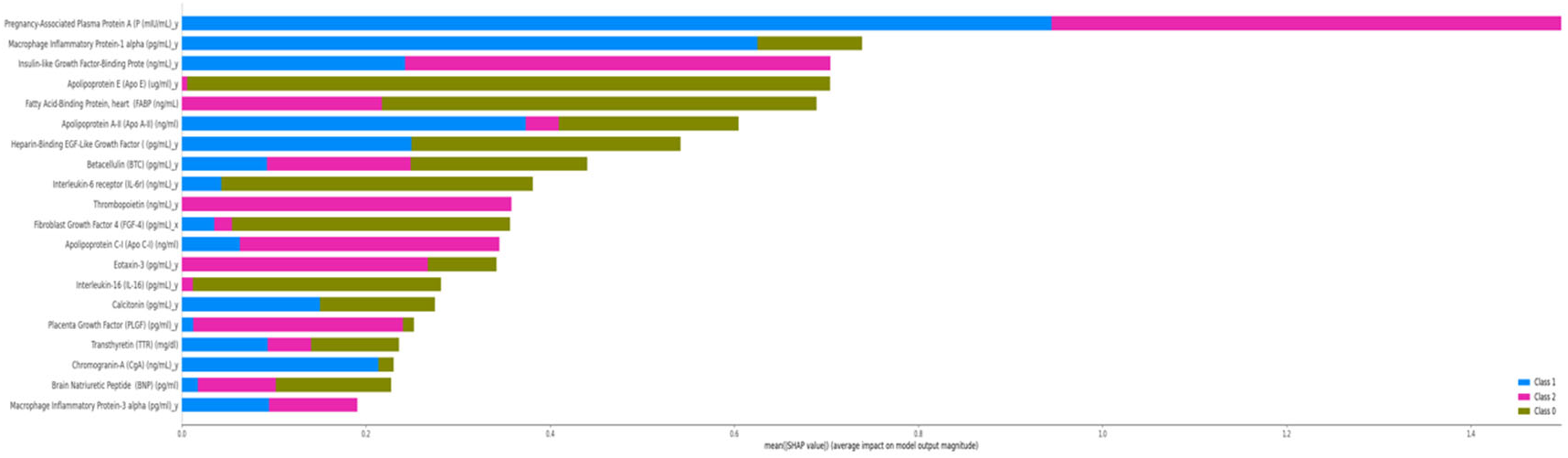
ADNI Summary Plot, Proteomic Features. The overlapping bars show the average SHAP values of the top proteomic features for each class.

**Figure 6:**
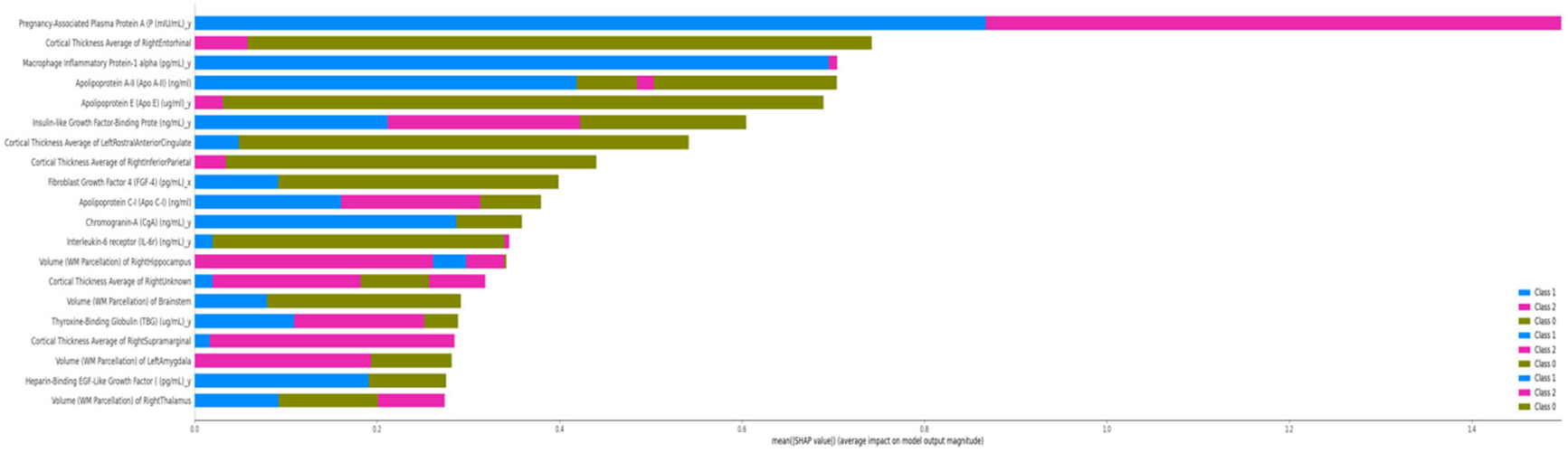
ADNI Summary Plot, All Features. The overlapping bars show the average SHAP values of the top imaging and proteomic features for each class.

**Table 1:**
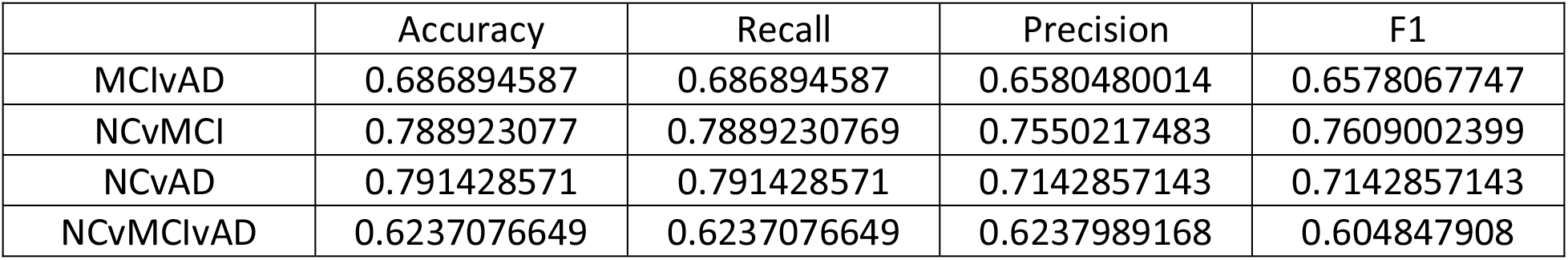
XGBoost performance on MRI Image Features Only. All scores are the average of 5-fold cross-validation. The recall, precision, and F1-scores for the trinary test (NCvMCIvAD) use a weighted average.

**Table 2:** XGBoost Performance on Proteomic Features Only. All scores are the average of 5-fold cross-validation. The recall, precision, and F1-Scores for the trinary test (NCvMCIvAD) use a weighted average.

|  | Accuracy | Recall | Precision | F1 |
| --- | --- | --- | --- | --- |
| MCivAD | 0.7612535613 | 0.7612535613 | 0.7602303998 | 0.7449376398 |
| NCvMCI | 0.8043076923 | 0.8043076923 | 0.8154784925 | 0.7897161275 |
| NCvAD | 0.7400000000 | 0.7400000000 | 0.7608008658 | 0.7339818077 |
| NCvMCivAD | 0.6295900178 | 0.6295900178 | 0.5937355078 | 0.5894027955 |

**Table 3:** XGBoost Performance on MRI and Proteomic Features Combined. All scores are the average of 5-fold cross-validation. The recall, precision, and F1-Scores for the trinary test (NCvMCIvAD) use a weighted average.

|  | Accuracy | Recall | Precision | F1 |
| --- | --- | --- | --- | --- |
| MCivAD | 0.8136752137 | 0.8136752137 | 0.8113215615 | 0.7942853606 |
| NCvMCI | 0.8043076923 | 0.8043076923 | 0.8154784925 | 0.7897161275 |
| NCvAD | 0.779047619 | 0.779047619 | 0.8011082251 | 0.7746031746 |
| NCvMCivAD | 0.6896613191 | 0.6896613191 | 0.6984263775 | 0.6676843769 |

Table 4 shows the results improvement from combining across all experiments.

**Table 4:** Results Improvement. This table shows how well combining the two datatypes improves performance. The performances are in terms of F1-Score.

| Problem | XGBoost Performance on Imaging Data | XGBoost Performance on Proteomic Data | XGBoost Performance on Combined Data | Performance Improvement |
| --- | --- | --- | --- | --- |
| ADNI MCivAD | 0.658 | 0.745 | 0.794 | 0.049 |
| ADNI NCvMCI | 0.761 | 0.790 | 0.790 | 0.000 |
| ADNI NCvAD | 0.714 | 0.734 | 0.775 | 0.041 |
| ADNI Trinary | 0.605 | 0.589 | 0.668 | 0.063 |

XGBoost displayed similar performance on both the imaging and proteomic datasets. For most of the experiments performed, combining the two kinds of data produced a synergistic increase in performance. The NCvAD experiment was an exception, showing no improvement when combining the two data sets. For the other three ADNI experiments, XGBoost produces an average improvement of 0.051 in terms of f1-score for the combined data.

The logistic regression model performed best out of all models tried for this experiment. Its performance is such that it may provide valuable information for diagnosis and prognosis of patients with suspected Alzheimer’s disease.

With its reasonable performance and self-validation by re-discovering previously known associations with AD, the XGBoost model has the potential to serve as a valuable diagnostic tool for patients with suspected AD.

## 4. Discussions

### 4.1 Performance of the different Models

#### XGBoost

Overall, XGBoost produced good results on the ADNI dataset, producing a weighted f1-score of 0.76 on the combined data. XGBoost produced good synergy when combining the data, leading to an f1-score improvement of 0.05.

#### Logistic Regression

Logistic regression tended to outperform XGBoost in this analysis. Though they performed equally on MRI data alone, logistic regression outperformed XGBoost on the proteomic and combined data, yielding a final f1-score of 0.82 compared to XGBoost’s 0.76. The logistic regression also showed more synergy between the datasets than XGBoost, improving by 0.07 when the datasets were combined.

#### K-Nearest Neighbors

Multiple values of K were tried for KNN. K=5 outperformed all other values of K on all datasets, although it still underperformed compared to XGBoost and logistic regression. The MRI dataset was less sensitive to changes in the value of K than the proteomic or combined datasets.

### 4.2 SHAP Feature Importance Analysis

The feature importance learned by XGBoost and reported using the SHAP utility matches up with expectations given current medical knowledge about AD. In MRI images, AD typically shows as non-specific generalized and focal atrophy of the brain [@Wolk]. Hippocampal and medial temporal atrophy are the most characteristic findings of AD but remain insufficient for diagnosis by imaging alone [@Wolk]. SHAP analysis of our XGBoost model showed that temporal lobe cortical thickness measurements were among the top features for the model using imaging features alone. Additionally, at least one hippocampal volume measurement appeared among top features for the model using imaging and proteomic features.

The importance of proteomic features shows a similar pattern. Apolipoproteins showed up frequently among the top features in both the model using proteomic features alone and the model using imaging and proteomic features. apolipoprotein E (APOE) and apolipoprotein ε4 (APOE ε4) have been previously studied as predictors for AD [@Wolk].

## Data Availability

All data produced in the present study are available upon reasonable request to the authors.

## ACKNOWLEDGMENTS

This research work was partially supported by the National Science Foundation EPSCOR DART grant, and the National Science Foundation with grant number 1452211, 1553680, and 1723529, National Institute of Health grant R01LM012601, as well as was partially supported by National Institute of Health grant from the National Institute of General Medical Sciences (P20GM103429).

Data collection and sharing for this project was funded by the Alzheimer’s Disease Neuroimaging Initiative (ADNI) (National Institutes of Health Grant U01 AG024904) and DOD ADNI (Department of Defense award number W81XWH-12-2-0012). ADNI is funded by the National Institute on Aging, the National Institute of Biomedical Imaging and Bioengineering, and through generous contributions from the following: AbbVie, Alzheimer’s Association; Alzheimer’s Drug Discovery Foundation; Araclon Biotech; BioClinica, Inc.; Biogen; Bristol-Myers Squibb Company; CereSpir, Inc.; Cogstate; Eisai Inc.; Elan Pharmaceuticals, Inc.; Eli Lilly and Company; EuroImmun; F. Hoffmann-La Roche Ltd and its affiliated company Genentech, Inc.; Fujirebio; GE Healthcare; IXICO Ltd.; Janssen Alzheimer Immunotherapy Research & Development, LLC.; Johnson & Johnson Pharmaceutical Research & Development LLC.; Lumosity; Lundbeck; Merck & Co., Inc.; Meso Scale Diagnostics, LLC.; NeuroRx Research; Neurotrack Technologies; Novartis Pharmaceuticals Corporation; Pfizer Inc.; Piramal Imaging; Servier; Takeda Pharmaceutical Company; and Transition Therapeutics. The Canadian Institutes of Health Research is providing funds to support ADNI clinical sites in Canada. Private sector contributions are facilitated by the Foundation for the National Institutes of Health (www.fnih.org). The grantee organization is the Northern California Institute for Research and Education, and the study is coordinated by the Alzheimer’s Therapeutic Research Institute at the University of Southern California. ADNI data are disseminated by the Laboratory for Neuro Imaging at the University of Southern California.

